# Temporary increase in circulating replication-competent latent HIV-infected resting CD4+ T cells after switch to an integrase inhibitor based antiretroviral regimen

**DOI:** 10.1101/2023.05.12.23289896

**Authors:** Roux-Cil Ferreira, Steven J. Reynolds, Adam A. Capoferri, Owen Baker, Erin E. Brown, Ethan Klock, Jernelle Miller, Jun Lai, Sharada Saraf, Charles Kirby, Briana Lynch, Jada Hackman, Sarah N. Gowanlock, Stephen Tomusange, Samiri Jamiru, Aggrey Anok, Taddeo Kityamuweesi, Paul Buule, Daniel Bruno, Craig Martens, Rebecca Rose, Susanna L. Lamers, Ronald M. Galiwango, Art F. Y. Poon, Thomas C. Quinn, Jessica L. Prodger, Andrew D. Redd

**Author notes:** Correspondence to: Andrew D. Redd, Johns Hopkins University; Rangos Building Room 540, 851 N Wolfe St, Baltimore MD, 21205. ADR and JLP contributed equally on this study.

## Abstract

The principal barrier to an HIV cure is the presence of a latent viral reservoir (LVR) made up primarily of latently infected resting CD4+ (rCD4) T-cells. Studies in the United States have shown that the LVR decays slowly (half-life=3.8 years), but this rate in African populations has been understudied. This study examined longitudinal changes in the inducible replication competent LVR (RC-LVR) of ART-suppressed Ugandans living with HIV (n=88) from 2015-2020 using the quantitative viral outgrowth assay, which measures infectious units per million (IUPM) rCD4 T-cells. In addition, outgrowth viruses were examined with site-directed next-generation sequencing to assess for possible ongoing viral evolution. During the study period (2018-19), Uganda instituted a nationwide rollout of first-line ART consisting of Dolutegravir (DTG) with two NRTI, which replaced the previous regimen that consisted of one NNRTI and the same two NRTI. Changes in the RC-LVR were analyzed using two versions of a novel Bayesian model that estimated the decay rate over time on ART as a single, linear rate (model A) or allowing for an inflection at time of DTG initiation (model B). Model A estimated the population-level slope of RC-LVR change as a non-significant positive increase. This positive slope was due to a temporary increase in the RC-LVR that occurred 0-12 months post-DTG initiation (p<0.0001). This was confirmed with model B, which estimated a significant decay pre-DTG initiation with a half-life of 7.7 years, but a significant positive slope post-DTG initiation leading to a transient estimated doubling-time of 8.1 years. There was no evidence of viral failure in the cohort, or consistent evolution in the outgrowth sequences associated with DTG initiation. These data suggest that either the initiation of DTG, or cessation of NNRTI use, is associated with a significant temporary increase in the circulating RC-LVR.

**Author Summary:** HIV is a largely incurable infection despite the use of highly successful antiretroviral drugs (ARV) due to the presence of a population of long-living resting CD4+ T cells, which can harbor a complete copy of the virus integrated into the host cell*’*s DNA. We examined changes in the levels of these cells, referred to as the latent viral reservoir, in a group of ARV-treated Ugandans living with HIV. During this examination, Uganda authorities switched the backbone drug used in ARV regimens to a different class of drug that blocks the ability of the virus to integrate into the cell*’*s DNA. We found that for approximately a year after this switch to the new drug, there was a temporary spike in the size of the latent viral reservoir despite the new drug continuing to completely suppress viral replication with no apparent adverse clinical effects.

## Introduction

HIV persists in individuals treated with fully suppressive antiretroviral therapy (ART) due in large part to the presence of a population of latently infected immune cells that are collectively referred to as the latent viral reservoir (LVR) [1]. The LVR is primarily made up of resting memory CD4+ (rCD4) T cells, which can be found in the circulating blood and tissues throughout the body [2]. While ART effectively blocks new cells from becoming infected in people living with HIV, it has no direct effect on the stably integrated proviruses that comprise the LVR. The LVR is maintained in the presence of ART through clonal expansion of latently infected cells due to a combination of homeostatic proliferation and antigenic stimulation, which together with the relatively long-lived nature of rCD4 T cells, creates an estimated half-life of the entire LVR of approximately 44 months [3,4]. This means that to naturally clear the replication competent LVR (RC-LVR) on standard ART alone would take an estimated 75 years or longer, essentially making HIV a life-long disease.

The vast majority of work examining the LVR has been focused on high-income countries, even though more than two-thirds of people living with HIV are African. Earlier work by our group demonstrated that the RC-LVR, as measured by the quantitative viral outgrowth assay (QVOA), was significantly lower in Ugandans than in North Americans [5]. One possible explanation for this difference could be that there might be a more rapid decay of the LVR in this Ugandan population. To examine this hypothesis, the previous study of Ugandans was extended to include longitudinal large blood draws (2015-2020), which were subsequently used for determination of changes in the size of the RC-LVR over time.

During this longitudinal study, Uganda medical authorities began a nationwide roll-out of integrase strand transfer inhibitor (INSTI) based ART in 2018, consisting of Dolutegravir (DTG), Tenofovir (TDF), and Lamivudine (3TC). This change provided an opportunity to examine any effects the switch from a non-nucleoside reverse transcriptase inhibitor (NNRTI) based regimen,the primary ART backbone prior to the switch, to one containing an INSTI, may have on the size of the LVR.

## Results

The population used for the longitudinal analysis of the RC-LVR has been described in detail previously [5,6]. Briefly, people living with HIV from the Rakai and Kyotera Districts of Uganda who were virally suppressed (viral load <40 copies/mL) for at least 18 months were recruited for annual large blood draws to examine the size and make-up of their LVR (Table 1). An initial group of participants were enrolled in 2015 (n=70), and were followed annually in 2016, 2017, 2019, and 2020 [5]. For this initial group, annual samples were not collected in 2018 due to a delay in extending the protocol beyond three initial time points. An additional group of participants (n=20) were enrolled in 2016 and followed annually in 2017, 2018, and 2019 [6]. For each study time point, large blood draws (180 mL) were collected and separated into peripheral blood mononuclear cells (PBMC) and plasma, which were both frozen and stored in liquid nitrogen and -80°C freezers, respectively. One participant was lost to follow-up without providing a valid QVOA, and another was later excluded from the analysis due to a loss of viral suppression during the study.

**Table 1.** Summary of study participants.

|  | Full cohort<br>(n=88) | Individuals with multiple<br>timepoints (n=69) |
| --- | --- | --- |
| Switched to DTG (n, %) | 73 (83%) | 63 (91%) |
| Female sex (n, %) | 56 (64%) | 45 (65%) |
| HIV-1 Subtype (n, %) |  |  |
| D | 49 (55.7%) | 40 (58.0%) |
| A1 | 16 (18.2%) | 13 (18.8%) |
| C | 2 (2.3%) | 1 (1.5%) |
| Recombinants | 16 (18.2%) | 14 (20.4%) |
| No subtype determined | 5 (5.7%) | 1 (1.5%) |
| Pre-ART viral load, log<sub>10</sub> copies/ml | 4.62 (2.91 - 6.57) | 4.65 (3.00 - 6.57) |
| Nadir CD4, cell/mm<sup>3</sup> | 185 (3 - 939) | 168 (3 - 690) |
| Age at baseline, years | 42.4 (26.6 - 59.6) | 43.1 (31.6 - 59.6) |
| BMI at baseline, kg/m<sup>2</sup> | 22.0 (15.8 - 34.2) | 21.7 (15.8 - 34.2) |
| ART duration at baseline, years | 7.7 (1.5 - 12.1) | 8.2 (2.2 - 12.1) |
| Observation time, years | - | 5.0 (1.0 - 5.3) |
| Pre-DTG | - | 3.3 (1.0 - 6.2) |
| Post-DTG | - | 1.8 (0.3 - 2.0) |
| Pre-DTG visits (mean) | 1.7 | 1.9 |
| 0 | 1 (1.1%) | 0 (0%) |
| 1 | 36 (40.9%) | 18 (26.1%) |
| 2 | 39 (44.3%) | 39 (56.5%) |
| 3 | 10 (11.4%) | 10 (14.5%) |
| 4 | 2 (2.3%) | 2 (2.9%) |
| Post-DTG visits (mean) | 1.2 | 1.5 |
| 0 | 24 (27.3%) | 6 (8.7%) |
| 1 | 26 (29.6%) | 25 (36.2%) |
| 2 | 38 (43.2%) | 38 (55.1%) |

Median and range presented, unless otherwise stated. DTG = Dolutegravir; ART = Antiretroviral therapy; BMI = Body mass index

Frozen PBMC were thawed and rCD4 T cells were isolated through negative selection and used for the QVOA to obtain an estimate of the infectious units per million (IUPM) rCD4 T cells using a maximum-likelihood based method, as described previously [7]. Any sample that experienced contamination during cell culture was excluded. PBMC samples that had insufficient CD4+ T cells isolated to warrant resting cell isolation were plated as bulk CD4 cells (n=20), and excluded from this analysis. The resulting rCD4 T cell QVOA results (n=254) were used for all subsequent analyses, and included samples from 88 participants (Table 1). Participants were majority female (63.6%), and had been on ART for a median of 7.7 years when the study began (range = 1.5 – 12.1). There was a range of time points per participant included in the study (n=19, 13, 20, 31, and 5 with one, two, three, four and five time points, respectively) with a median of three time points. Of the 254 QVOA measurements included, 152 were obtained prior to DTG-initiation (59.8%). Temporal changes in IUPM estimates for all participants were visualized by time on ART and time since DTG-initiation (Figure 1A and B).

**Figure 1:**
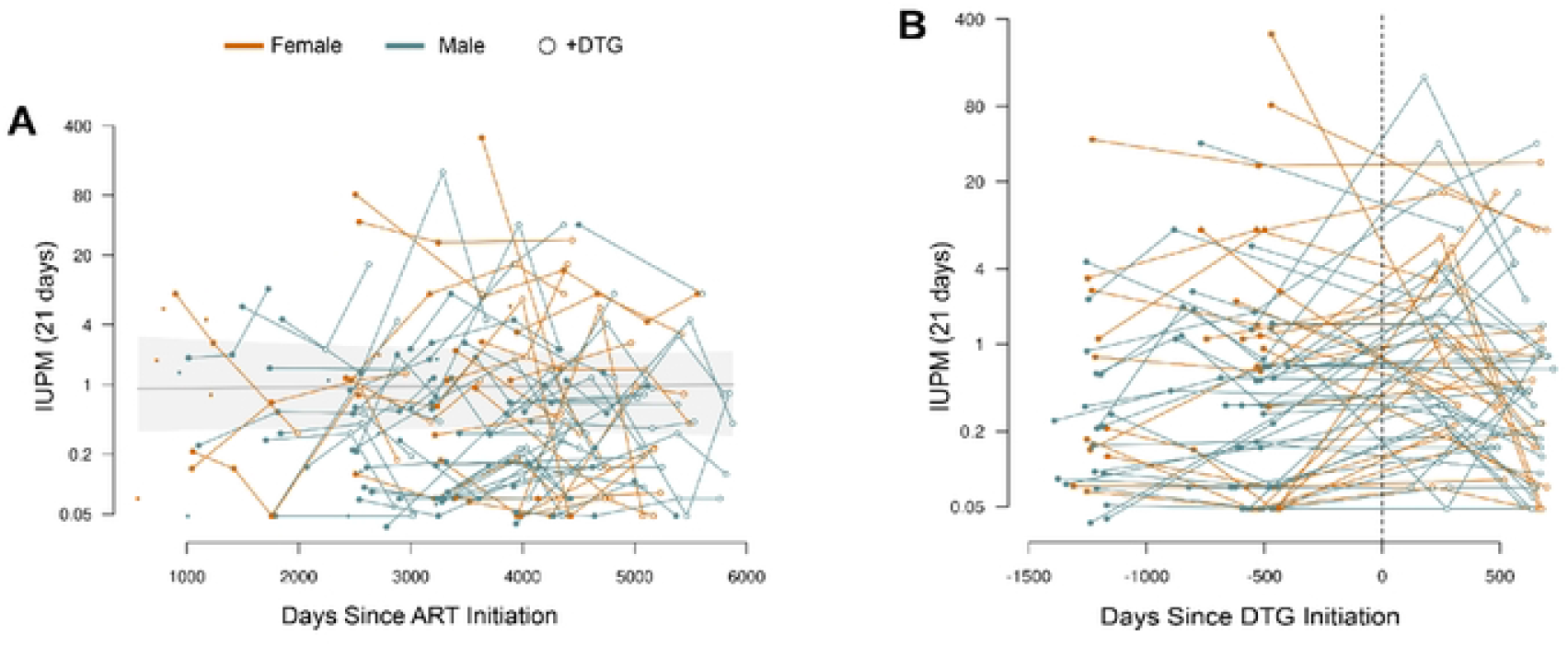
Change in Infectious units per million (IUPM) over time. Data are maximum likelihood estimates (MLEs) of IUPM basedon quantitative viral outgrowth assays (QVOA, 21 days outgrowth). Participants are aligned based on time of ART initiation **(A)** or time of dolutegravir (DTG) initiation **(B)**. Open points correspond to samples taken after the participant switched to a DTG-containing drug regimen. Points representing participants with only one sample are reduced in size. Longitudinal samples from the same study participant are connected by line segments and coloured withrespect to biological sex (seecolour legend). The grey line on (A) is theregression trend for the population-level posterior median values of IUPM over time (mixed-effects logistic regression model); light grey region is the 95% highest posterior density.

QVOA well-level data was used in two Bayesian hierarchical linear models (A and B) to examine the longitudinal change in the RC-LVR of the cohort. Both models used time since ART initiation as the time variable. However, model B incorporated an inflection in the model at the time of DTG initiation, allowing for different rates of change pre- and post-DTG. In addition to the longitudinal change, the models also examined the effects of biological sex, and were fit using the full dataset (n=88) and a subset that included only individuals with multiple time points (n=69; Table 1). These hierarchical models included both individual-level and cohort-level parameters (i.e. hyperparameters). Individual-level parameters estimated the linear longitudinal changes in the RC-LVR for each participant while the cohort-level parameters related individual-level parameters between the participants within the cohort. The hierarchical structure facilitates the interpretation of cohort-level features despite individual-level differences between the participants.

Using the full cohort dataset, the single individual-level rate model (model A) estimated that the size of the RC-LVR for the group was increasing in a non-significant manner (doubling time = 106.5 years; IQR = -41.4 – 23.4; Figure 2B). The individual participant estimated slopes were highly variable, ranging from significant decreases to significant increases. Female sex was associated with a smaller RC-LVR, supporting previous findings from this same population (median *β*_*sex*_= -1.23; IQR = -1.6 – -0.9, Figure 2C) [6].

**Figure 2.**
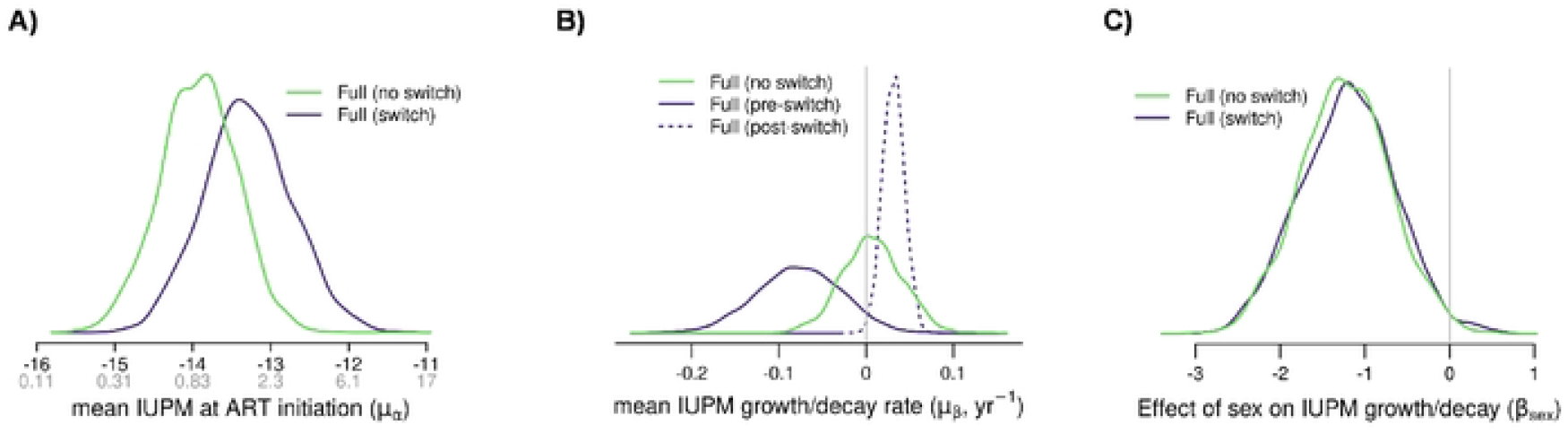
Posterior densities of logistic regression model parameters for the full cohort analysis. The curves (green for model #1 (no-switch] and purple for model #2 (switch]) represents the posterior density for the mean IUPM at initiation (A), the mean IUPM change (B), and the effect of biological sex (C) as indicated by the axis label. “Switch” indicates that the regression model #2 was used, Which includes a fixed effect for the rate of IUPM growth/decay before (solid line) and after (dotted line) DTG initiation. Standard deviations for the posterior densities are shown in the supplementary (Figure S2)

Model B was subsequently used to examine changes in RC-LVR on a cohort and individual-level for both pre- and post-DTG initiation. By allowing for an inflection point in the linear estimate of the slope of the change in IUPM over time, model B estimated that the size of the RC-LVR was significantly decreasing pre-DTG initiation (half-life = 9.0 years; IQR = 6.3 – 15.9; Figure 2B). Conversely, post-DTG initiation, the slope of the change in the RC-LVR was significantly increasing (doubling time = 9.5; IQR = 7.5 – 13.2; Figure 2B). Sex had a similar effect in model B as in model A, with female sex associated with a significantly smaller reservoir (*β*_*sex*_= -1.2). Similar to model A, the individual participant slopes were highly variable, ranging from significant decreases to significant increases pre- and post-DTG initiation. Similar findings were found for both models using the cohort with multiple time points (Supplemental Figures S1, S2, and S3)

Model A was then used to examine if the maximum likelihood IUPM estimates were significantly different from those predicted by the model, for each timepoint of each individual (Figure 3). Values were deemed significantly different if both the estimated point estimates were not included in the 95% confidence intervals of the maximum likelihood estimates or the highest density intervals for that given time point. Of the 240 individual time points that were tested for outgrowth at day 21 in culture, 16 (6.7%) and 4 (1.7%), were significantly higher or lower than the model predictions, respectively (Table 2). When examining the distribution of these outlying observations according to three time periods, pre-DTG, 0–1 year post-DTG, and >1 year post-DTG), there was a significantly higher percentage of individual time points with higher observed IUPM in the first year after DTG initiation, with 25% (11/44) of time points in that group being higher than what was estimated by the model (p<0.0001, chi-square test; Table 2). The percentage of significantly higher maximum likelihood IUPM estimates pre-DTG, and post-DTG (>1 year) were 2.2% (3/139) and 3.5% (2/57), respectively (Table 2). This analysis was also performed stratified by subtype of the infecting strain (determined by outgrowth sequencing of *pol* and *env*) and similar patterns were seen in subtype D (p<0.0001; Table S1). There were not enough significantly different time points observed in the subtype A or recombinants and other categories to examine these groups for significant differences.

**Table 2.** Significant differences between maximum likelihood and model A estimated IUPM
measurements by time point pre- and post-DTG initiation.

| Time window | Observed > Model<br>n (%) | Observed < Model<br>n (%) | Total<br>(n=240) |
| --- | --- | --- | --- |
| Pre-DTG initiation | 3 (2.2%) | 1 (0.7%) | 139 |
| Post-DTG (0-1 year) | 11 (25%) | 1 (2.3%) | 44 |
| Post-DTG (>1 year) | 2 (3.5%) | 2 (3.5%) | 57 |

\*chi-square significance $p < 0.0001$ , Full cohort dataset analysis.

**Figure 3.**
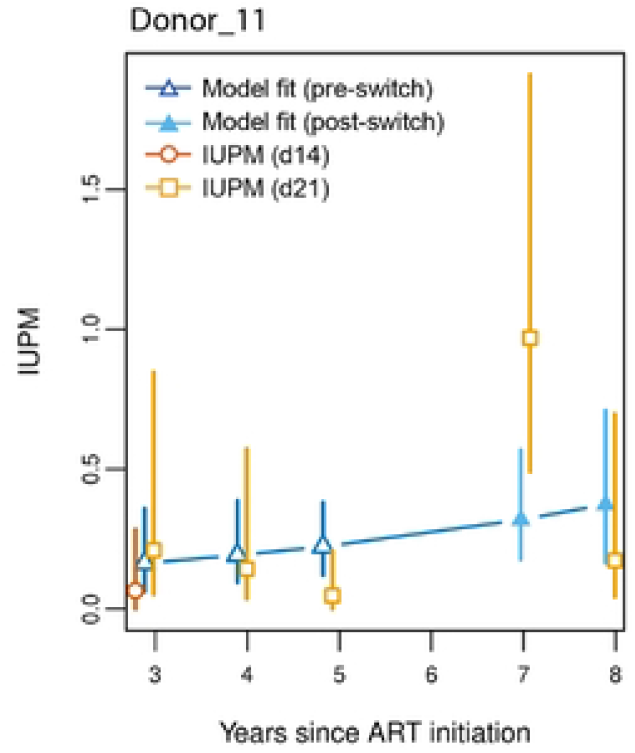
Representative graph of Donor 11*’*s observed day 14 (maroon) andday 21 (gold) IUPM measurements vs model estimated values for pre-DTG (dark blue opensymbols) and post-DTG initiation (light blue closed symbols) for one subject. 95% confidence intervals (Cl) for observed and model-estimatedvalues shown. Significance was determined if the day 21 point estimates for an IUPM and model-estimated value werenot included in the Cl of its partner measurement from the same time point.

One possible explanation for the apparent increase in the RC-LVR in the time points immediately post-DTG initiation would be a short-term loss of viral control during this period. Viral load measurements for all participants (n=88) were examined for any failures during the study period, but no cases of failure were documented (viral load > 200 copies/mL). In addition, a portion of wells where viral outgrowth was detected in the QVOA were sequenced using a site-directed next-generation sequencing (NGS) assay for the reverse-transcriptase (RT) portion of the *pol* gene and the gp41 region of the *env* gene, as previously described [8]. The assay can distinguish if a positive well contains one or more viral outgrowth species, and allows for phylogenetic analysis of these outgrowth viruses to examine if there is any evidence of ongoing evolution in the RC-LVR post-DTG initiation. Subjects with sufficient longitudinal NGS outgrowth data derived from rCD4 QVOA (n=41) were examined for any signs of continuing evolution by examining change in pairwise distance overtime using a linear regression model as previously described [9]. Of the subjects with available NGS data from pre- and post-DTG initiation, there was a median of three time points available per individual (range= 2-5), and a total of 909 wells were sequenced (Figure 4). There were eight individuals (19.5%) with significant changes in their pairwise distances in both *pol* and *env* over time (Table 3). There were 11 (26.8%) individuals who had a significant increase in pairwise distance of either *pol* or *env*, but not the other, and 22 individuals (53.7%) had no significant changes in either region. It should be noted that due to the large number of comparisons that are generated in pairwise distance measurements of this nature, even small changes, may be found to be significant. Therefore, estimates of pairwise changes for individuals with significant increases in their IUPM 0-12 months post-DTG initiation were compared to those who did not have significant increases during that time. Individuals with increased IUPM measurements were not more likely to have significantly changed pairwise distances (p=0.33, chi-square test), which supports the viral load analysis that the increases seen in the IUPM were not due to underlying ongoing viral replication.

**Table 3.**
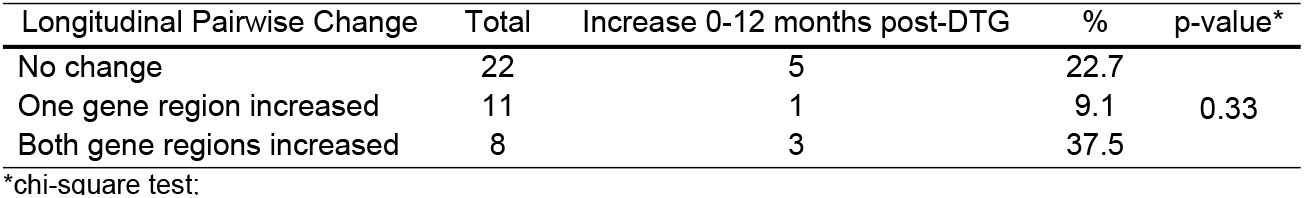
Change in longitudinal pairwise distance in pol and env vs significant increase in IUPM 0-12 months post-DTG

| Longitudinal Pairwise Change | Total | Increase 0-12 months post-DTG | % | p-value* |
| --- | --- | --- | --- | --- |
| No change | 22 | 5 | 22.7 | 0.33 |
| One gene region increased | 11 | 1 | 9.1 |  |
| Both gene regions increased | 8 | 3 | 37.5 |  |

\*chi-square test;

**Figure 4:**
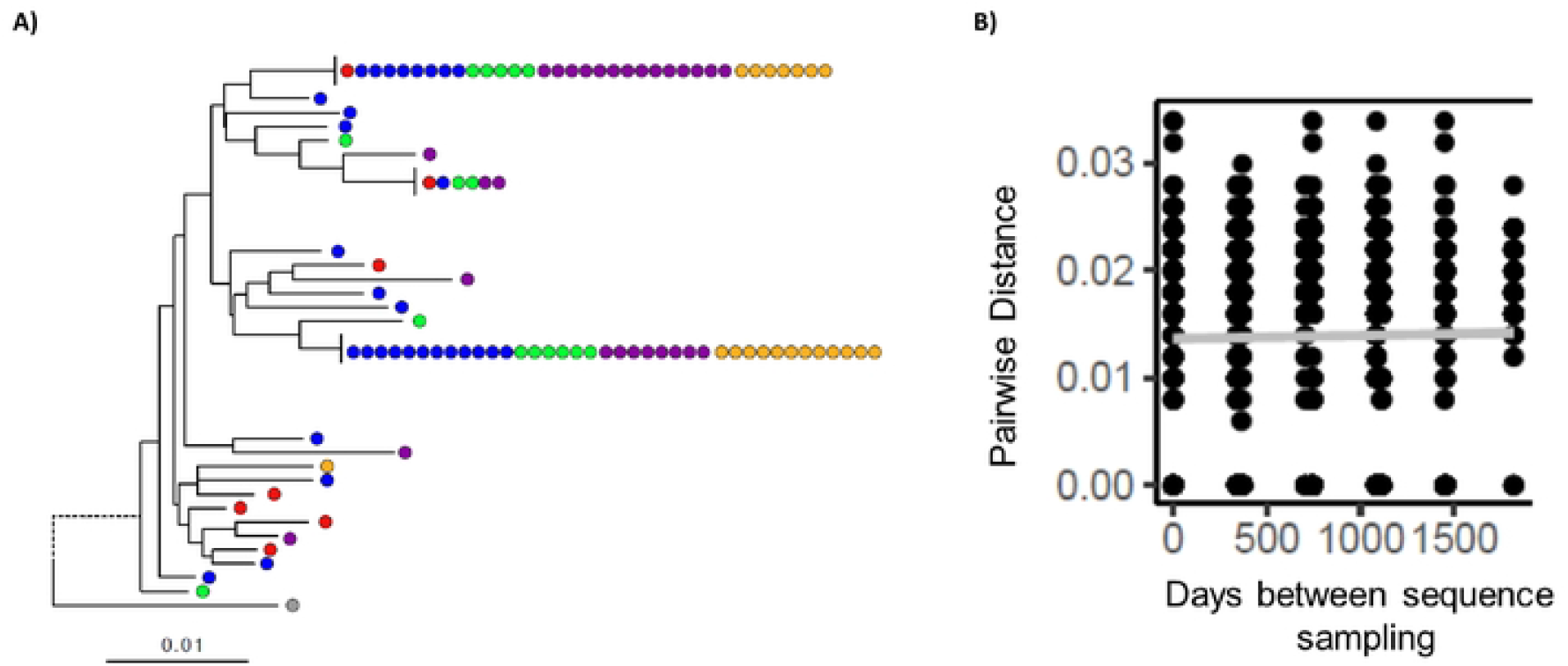
Representative neighbor-joining phylogenetic tree of viral pol sequences derived from next-generation sequencing of viral outgrowth viruses from Donor 17, who experienced a significant increase post-DTG initiation. Sequences derived from outgrowth viruses from a specific well are shownas individual circles, and wereisolated frompre-DTG switch (baseline (0, red) and years 1 (blue), 2 (green)) and post-DTG switch time points [years 4 (purple), and 5 (gold)), respectively (A). A representative sequence for HIV-1 subtypes B (HXB2) is shownin grey. Genetic distance is indicated by scale at the bottom. These data wereusedto calculate the pairwise distance over time for Donor 17 and examined for significant changes by linear regression (B; r^2^=0.00. p=0.181).

## Discussion

These data are the first detailed longitudinal analysis of the LVR in an African population, and identified a possible temporary effect of DTG-treatment initiation on the size of the RC-LVR in this population. The initial observation that the RC-LVR was slightly increasing was the opposite of the expected observation given the previous findings of a smaller RC-LVR in this Ugandan population versus comparable North Americans [5]. However, given that the majority of individual time points that were significantly higher than the model prediction were found immediately post-DTG initiation, and that when an inflection point was allowed in model B there was a significantly negative slope pre-DTG, suggests that the natural pattern of the RC-LVR in this population was declining, if not for the temporary increase seen post-DTG initiation. In addition, the modeled rate of decline pre-DTG switch corresponds to a half-life of nine years, which is slower than previous estimates using QVOA derived IUPM data for North American populations [3,4]. In the North American population, the half-life of overall LVR decline was approximately 3.7 years, which would appear to be significantly faster than the rate of decline observed here. However, there are some important caveats that make a direct comparison difficult. First, the Ugandan population was primarily female, while HIV latency studies done in North America and Europe include predominantly male cohorts. The current analysis and other studies suggest that there are important sex-based differences in the RC-LVR [6,10,11]. In addition, most individuals in our study were on ART for >7 years prior to study enrollment, and some studies have recently found a tri-modal decay pattern of intact proviral species, with the third phase that consists of very slow proviral decay starting approximately seven years after ART initiation (half-life=18.7 years). It is possible that the relatively longer half-life observed here reflects this third, very slow phase of decay [12].

The finding of an increase in the RC-LVR for approximately one-year post-DTG initiation was a surprising observation, and the return to lower levels after one year suggests that this increase was temporary. Interestingly, other research studies have also found short-term effects related to DTG-initiation in people living with HIV. In particular, the RESPOND study, a large analysis of Europeans and Australians with HIV, found that the risk of a cardiovascular disease event almost doubled for the first six months in individuals who initiated an INSTI, and that this risk decreased over the next 18 months to return to normal [13]. In addition, in a small exploratory study of individuals who were virally suppressed on non-INSTI regimens it was found that there are subtle shifts in the overall make-up of the rCD4 cell population 48 weeks post-DTG initiation that lead to a decrease in total HIV DNA levels in effector memory cells [14]. This same study found that there is also a temporary decrease in viral diversity post-DTG initiation [15]. In all of these cases, the effects observed were temporary, similar to the increase in RC-LVR seen here. It should be noted that the INDOOR study found no changes in total HIV DNA or cell-associated RNA post-DTG initiation in a small group of virally suppressed individuals on protease-inhibitor containing regiments [16]. This highlights an important caveat of our findings since the majority of the Ugandans were on NNRTI-based regimens pre-DTG initiation. It is possible that it is the removal of the NNRTI and not the addition of DTG that is causing the effect seen here. In addition, with only one follow-up time point post the temporary increase in the RC-LVR, it is unclear what the long-term effects on the LVR will be in these individuals, but this work is ongoing.

One possible explanation for this temporary increase in the RC-LVR is a temporary change in the circulating immune cell make-up post-DTG initiation. This is supported by an analysis of the relatively large-scale SWORD 1 and 2 studies, where it was found that in virally-suppressed individuals who switched to a two-drug regimen of DTG and an NNRTI (Rilpivirine), there was temporary increase of soluble CD14 (sCD14), which is a marker of monocyte activation [17]. However, the INDOOR study did not see changes in cytokines, and a smaller study of individuals switching from an NNRTI (Efavirenz) to DTG found a decrease in sCD14 eight weeks post-switch [16,18]. Understanding the possible immunological mechanism behind the temporary increase in the RC-LVR seen here is currently being explored.

A limitation of the current study is our inability to use the intact proviral DNA assay (IPDA) in the Ugandan cohort, because that assay has only been validated for HIV subtype B [19]. Another limitation is that the QVOA has a high level of inter- and intra-subject variability. The effect of this variability was mitigated by modeling well-level outgrowth data, as well as using the group and individual level data together. In addition, the size of this cohort helps to mitigate QVOA variability. While there were no documented cases of loss of viral suppression (outside the excluded individual), this prospective study was not designed to capture the effect of DTG initiation, and therefore the timing between DTG initiation and the subsequent viral load/QVOA measurements were not measured directly after post-DTG initiation. However, there was no consistent evidence of viral evolution in individuals who experienced significant increases in their RC-LVR, suggesting that low-level viral failure is not contributing to this finding. Another limitation is that a portion of the baseline QVOA assays were measured only at day 14 of viral outgrowth (based on protocols existing at that time) and not continued to day 21. However, this was included and accounted for in both models. Lastly, 19 individuals in the study only contributed one RC-LVR measurement to the analysis. This was due to missed sample collections, contamination, or, excluding any sample that used bulk CD4+ T-cells for the QVOA. To address this, the analysis was repeated using only people who contributed more than one time point, with virtually identical results.

It will be critical to examine if the temporary increase seen in these Ugandans is found in other populations of suppressed individuals who switched to DTG or another INSTI. In addition, it will be important to examine if this change is due to an overall increase in the total LVR or a shifting of the latently-infected rCD4 T cell population to a more inducible phenotype for 0-12 months post-DTG switch, which could have important implications for possible HIV cure strategies.

## Materials and Methods

### Study Population

The details of the population examined for this analysis have been discussed in detail previously [5,6]. Briefly, people living with HIV-1 from the Rakai and Kyotera Districts of Uganda who were virally suppressed (viral load <40 copies/mL) for at least 18 months were recruited for annual large blood draws (∼180 mL) to examine the size and make-up of their LVR (Table 1). An initial group of participants were enrolled in 2015 (n=70), and were followed annually in 2016, 2017, 2019, and 2020 [5]. An additional group of participants with estimated dates of seroconversion (n=20) were later enrolled in the study in 2016 and followed annually in 2017, 2018, and 2019 [6]. For each study time point, blood was collected and separated into peripheral blood mononuclear cells (PBMC) and plasma, which were both stored for later analysis. All participants underwent a clinical examination, viral load, and CD4+ cell count at each study visit. One participant was excluded from the analysis due to a loss of viral suppression during the study due to being incarcerated.

All participants provided written informed consent, and ethical approval was obtained from the Institutional Review Boards at National Institutes of Allergy and Infectious Diseases (NCT02154035), Uganda Virus Research Institute, Uganda National Council for Science and Technology, and Johns Hopkins University.

### Quantitative viral outgrowth assay

QVOA was performed as previously described [5,6]. Briefly, frozen PBMC were thawed and CD4+ cells were isolated using negative selecting bead purification (CD4+ T-cell Isolation Kit II, Miltenyi). The number of viable CD4+ T cells were counted and if the sample contained sufficient numbers rCD4 cells were isolated using negative selecting bead purification (anti-CD25, anti-biotin MicroBeads, anti-CD69 MicroBead Kit II, and anti–HLA-DR MicroBeads; Miltenyi). The resulting rCD4 cells were stimulated with Phytohaemagglutinin (PHA) and *γ*-irradiated PBMC and co-cultured with MOLT-4 cells transfected with CCR5 and naturally expressing CD4 and CXCR4 (MOLT4/CCR5; National Institutes of Health AIDS reagent program). The co-culture supernatants were tested for presence of HIV p24 by ELISA (PerkinElmer) after 14 or 21 days, to indicate outgrowth of replication-competent provirus [20]. For 2015 samples, all rCD4 cells were plated in a limiting dilution as previously described [20]. A portion of 2015 QVOA were tested for outgrowth viruses at 14 days, which was included and considered in both of the models. All time points collected after 2015, were measured at 21 days, with some also being tested at 14 days. In addition, QVOA for post-2015 samples were plated with a standard limited diluting plating strategy of approximately 14.5×106 total rCD4 cells.

### Next-generation sequencing of outgrowth viruses

Outgrowth viral sequences from p24 positive wells were obtained as previously described [8]. Briefly, viral RNA was isolated from culture supernants of p24 positive wells, next-generation site-directed sequencing libraries were created for reverse transcriptase (*pol*; HXB2 position 2723–3225) and gp41 (*env*; HXB2 position 7938–8256), and sequenced (Illumina Inc, San Diego CA). Identical sequence reads were combined and the prominent strains (sequences with >2.5% of the total sequence reads analyzed for a given sample) were identified, cleaned of possible intra-well recombinants, and aligned for all time points from a given person (Figure 4) [8,21]. These *pol* and *env* sequences were used to determine HIV subtype by phylogenetic analysis, and the alignments were used to calculate the amount of change in the pairwise distance overtime using a linear regression model as previously described [9]. Briefly, raw pairwise differences between all sequences in the alignment were calculated and compared to the time between the two sequences. These values were then examined using a linear regression to determine if there was a significant association overtime. The R^2^ and p-values were collected for all individuals who had sequence data available from two or more time points, and compared between individuals who experienced a significant increase in their IUPM 0-12 months post-DTG initiation and everyone else with available sequence data. Differences were examined by chi-square analysis.

### Data analysis

Viral load and QVOA data were linked to ART regimens and visit dates and merged into a data frame in the R statistical computing environment. We assume the number of p24 positive outgrowth wells after 21 days is a binomial outcome with probability P=1−exp(−*n*_*i*_*λ*), where *n*_*i*_ is the number of cells in the *i*-th well and *λ* is the IUPM. This outcome is further partitioned into wells that do or do not have positive outgrowth after 14 days with a probability *p*_14d_. Assuming wells with positive outgrowth at 14d are always positive at 21d, the QVOA results can modeled as multinomial outcomes with probabilities *p* (*p*_14d_) (outgrowth at 14d and 21d), *p* (1 – *p*_14d_) (outgrowth at 21d only), and (1 – *p*) (no outgrowth), respectively. Given this formula and outgrowth data, we used the non-linear minimization algorithm in R (function *nlm*) to estimate the maximum likelihood IUPM for each subject and timepoint. We calculated 95% confidence intervals from the Hessian matrix for each estimate. Next, we fit a mixed effects logistic regression to these data:

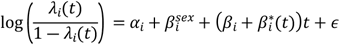

where *λ*_*i*_(*t*) is the IUPM for the *i-*th subject at *t* days after ART initiation, subject-specific intercept *α*_*i*_ and slope *β*_*i*_ are normally distributed random effects, 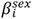 is the categorical fixed effect of sex, 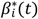 is the change in IUPM growth/decay rate upon switching to an integrase inhibitor-containing regimen, and *ε* is the error term. We implemented this model in the RStan package, which provides an R interface to Stan, a Bayesian statistical programming language (https://mc-stan.org), to generate a posterior sample of the model parameters. We ran four replicate chain samples for a warm-up period of 1,000 steps and then 3,000 steps, recording every 4 steps. Further details about the Stan analysis are provided as Supplementary Material. In addition, we ran posterior predictive checks by simulating QVOA data, *i*.*e*., number of positive wells at 14 and 21 days, from the multinomial distribution parameterized from the Stan outputs.

## Data Availability

All data produced in the present study are available upon reasonable request to the authors

## Acknowledgements

The authors would like to thank the RHSP study staff and the study participants.

